# Prevalence and risk factors associated with Anxiety Symptoms and Disorders Among Chronic Kidney Disease patients – A Systematic Review and Meta-analysis of studies

**DOI:** 10.1101/2020.06.03.20121798

**Authors:** Caleb Weihao Huang, Poh Hui Wee, Lian Leng Low, Ying Leng Agnes Koong, Htay Htay, Qiao Fan, Wai Yin Marjorie Foo, Jun Jie Benjamin Seng

**Author notes:** Details of corresponding author Name: Jun Jie Benjamin SENG (Mr) Address: 8 College Road, Singapore 169857. Authors contributed equally to this work (Caleb Weihao Huang, Jun Jie Benjamin Seng).

## Abstract

**Background:** Anxiety is associated with poor health outcomes among chronic kidney disease (CKD) patients. This review summarizes the prevalence and risk factors associated with anxiety symptoms and disorders among CKD patients.

**Methods:** Articles evaluating the prevalence and risk factors associated with anxiety symptoms and disorders among CKD patients, as diagnosed via DSM 4^th^ or 5^th^ edition criteria, clinical interviews or validated questionnaires, were searched in Medline®, Embase®, PsychINFO® and CINAHL®. Using random-effects meta-analyses, prevalence of anxiety was estimated. A narrative review on the risk factors associated with anxiety was presented.

**Results:** From 4941 articles, 61 studies were included. Pooled prevalence of anxiety disorders (9 studies, n=1071) among CKD patients across studies was 18.9% while that of anxiety symptoms (52 studies, n=10,739) was 42.8%. Across continents, prevalence of anxiety symptoms was highest in Europe and Asia. Between pre-dialysis and dialysis patients, prevalence of anxiety symptoms was statistically comparable at 30.5% and 42.1% respectively. Most commonly studied risk factors associated with anxiety were female gender, increased age, concomitant depression, and increased comorbidities.

**Conclusion:** Given the high prevalence of anxiety disorders and symptoms, there is a need for developing clinical guidelines on anxiety screening among CKD patients, facilitating early identification of at-risk patients.

## Introduction

Chronic kidney disease (CKD) is a prevalent public health problem which afflicts 11–13% of the world’s population (1) and is associated with increased morbidity (2) and healthcare costs (3). As the disease progresses, CKD culminates in significant somatic symptoms, reduced health-related quality of life (HrQOL) and lifestyle limitations (4, 5). Unsurprisingly, CKD patients experience and suffer from significant psychological distress (6).

Anxiety is described as the feeling of fear, uncertainty, helplessness and apprehension that an individual encounters when anticipating a threatening situation (6). Conversely, pathological anxiety disorders differ from anxiety symptoms as they tend to be pervasive, span over a minimum of six months and have a propensity to worsen if left untreated. Anxiety disorders encompass a range of psychopathologies such as general anxiety disorder (GAD), panic disorder and social anxiety disorder (7). While there are multiple instruments validated for the assessment of anxiety such as the Hospital Anxiety Depression Scale (HADS), these symptoms are commonly overlooked in CKD patients as they may not display overt symptoms.

Anxiety has important implications on clinical and psychological outcomes among CKD patients. Across the spectrum of CKD ranging from pre-dialysis to end-stage renal disease (ESRD), anxiety symptoms have been consistently associated with significant impairments in patients’ HrQOL (8–10) and non-adherence to medical treatment (11, 12). It has also been associated with increased morbidity and mortality in CKD patients. A study by Loosman et al. showed that pre-dialysis CKD patients with anxiety had a 60% higher risk of mortality, hospitalization, or requiring initiation of dialysis compared to patients without anxiety (13).

Notably, anxiety was highlighted as a key research area in the Kidney Disease: Improving Global Outcomes (KDIGO) Controversies Conference in Supportive Care (14). While multiple studies have evaluated the relationship between anxiety and CKD, the prevalence of anxiety varies widely due to inter-study differences in patient populations, instruments used and study designs. A review by Murtagh et al. estimated that 38% of ESRD patients experienced anxiety symptoms (15). However, anxiety only formed a small component of CKD-related symptoms investigated in this review, which was conducted in 2007. Other reviews in literature are limited to narrative summaries performed for anxiety in select CKD populations (e.g. hemodialysis patients) (16) or pertaining to the challenges and management of anxiety. Additionally, there are no reviews which have evaluated the prevalence of anxiety disorders among CKD patients, to our best knowledge. To understand the unique needs of CKD patients with anxiety and guide future research priorities, summarizing the prevalence and risk factors associated with anxiety disorders and symptoms among adult CKD patients is essential. Hence, this review aims to summarize the prevalence and risk factors associated with anxiety symptoms and disorders among CKD patients.

## Methodology

### Search terms

We conducted a literature search for relevant articles in Medline®, Embase®, PsychINFO® and CINAHL®, in accordance to Preferred Reporting Items for Systematic Reviews and Meta-Analyses (PRISMA) checklist. The search terms used encompassed subtypes of anxiety disorders within the Diagnostic and Statistical Manual of Mental Disorders 4^th^ edition (DSM-IV) or 5^th^ edition (DSM-V), CKD related terms and anxiety symptoms. They were derived and adapted from systematic reviews which examined the prevalence of anxiety symptoms and disorders in other patient populations (17–19). The full search strategy is listed in Supplementary File 1. The literature review was current as of May 2020.

### Review and inclusion of articles, and data extraction

The review and inclusion of articles were performed by two independent reviewers (CW Huang and PH Wee). All citations were evaluated initially for relevance by their title and abstract. Thereafter, the full-text articles of potentially relevant articles were retrieved and assessed. Discussion was made with a third independent reviewer (JJB Seng) to reach a consensus for all disagreements. Full-text, peer-reviewed, original articles published in English which examined the prevalence of anxiety disorders or symptoms in observational studies among adult CKD patients (age > 18 years old), as assessed using the DSM-IV or DSM-V criteria in clinical interviews or validated anxiety rating scale questionnaires were included. Case reports, case series, unrelated systematic reviews, meta-analyses and studies involving kidney transplant patients were excluded. For studies that had multiple papers examining anxiety using same patient dataset, only the study with the complete dataset was included to avoid duplication and skewing of results. Hand-searching within the reference lists of included articles was further performed to identify additional relevant articles.

Data were collected using a standardized data extraction form. Studies were segregated into those whose participants had clinically diagnosed anxiety disorders and those whose deemed to have anxiety symptoms based on anxiety rating instruments. Risk factors associated with anxiety symptoms and disorders were categorized into four main domains (patient-related, medical-condition related, therapy-related and psychosocial-related). In view of the limited number of studies which examined anxiety disorders, the risk factors analyses for anxiety disorders and symptoms were combined.

### Quality assessment of studies

The Quality Assessment Tool for Observational, Cohort and Cross-Sectional Studies by National Health, Lung and Blood Institute was used to evaluate the methodological quality of studies (14). Articles were scored as low, moderate and high risk of bias by two independent reviewers (CW Huang and PH Wee) according to responses in the checklist. Should insufficient information permit scoring of an item, original authors of the study were contacted for clarification. If authors were not contactable, the item was rated as high risk of bias (20).

### Statistical Analyses

Meta-analyses were performed using Stata software, version 14.0. To evaluate the pooled prevalence rate of anxiety symptoms and disorders, random-effects model was used due to heterogeneity of included studies. Heterogeneity across studies was evaluated using the I^2^ statistic (21). Subgroup analyses were only performed for studies which examined anxiety symptoms due to small number of studies which evaluated the prevalence of anxiety disorders. The analyses were stratified by patient population (pre-dialysis, dialysis and both), continent of study, and type of anxiety instrument.

## Results

Figure 1 shows the flowchart for the inclusion of articles. Of the 4941 citations retrieved, 61 studies were included in this review. Overall, the risk of bias was low in 42(68.9%) and moderate in 19 (31.1%) of the studies (Supplementary File 2). Majority of the study designs were cross-sectional in nature (n = 51, 83.6%) while eight studies were prospective cohort studies (n = 10, 16.4%).

**Figure 1:**
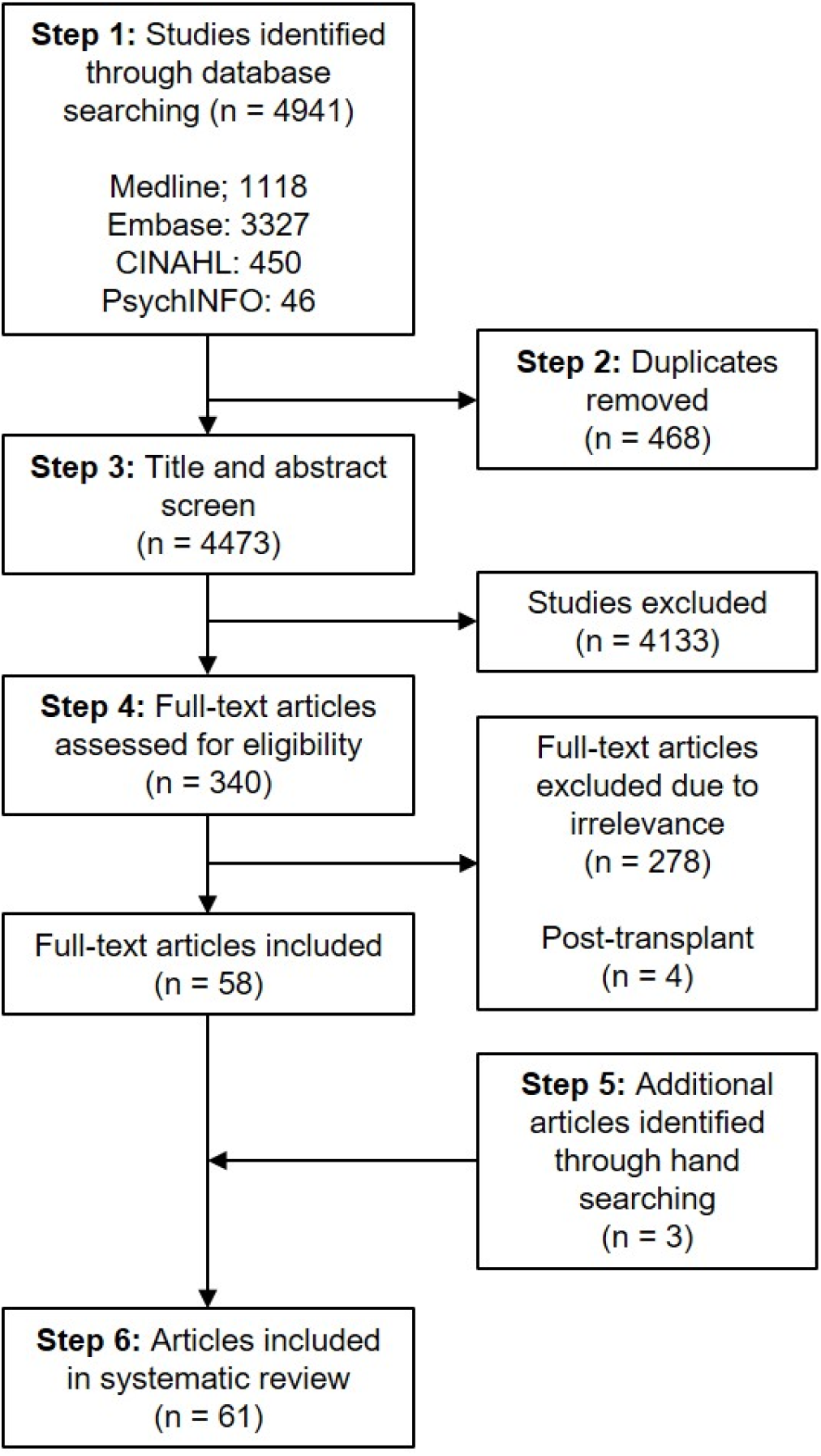
Flow chart of studies retrieved from database searching and the number of citations included and excluded from the systematic review

Table 1 summarizes details of the patient populations and tools used for the assessment of anxiety within the included studies. Detailed information regarding each study is reported in Supplementary File 3.

**Table 1.** Overview of included studies (n = 61)

| Study (Year) | Country | Study Design | No. of patients | Mean age (years) | Gender, [Men (%)] | Stage of CKD | Modality of dialysis (Duration) | Instrument used for anxiety symptom or disorder assessment | Mode of administration of interview or instrument | Subtypes of Anxiety Disorders studied |
| --- | --- | --- | --- | --- | --- | --- | --- | --- | --- | --- |
| **Anxiety disorders** |  |  |  |  |  |  |  |  |  |  |
| Oyekcin et al. (2012) | Turkey | Cross-sectional | 90 | 37.6 (HD)<br>36.9 (PD) | 61.2 (HD)<br>50 (PD) | Stage 5D | HD or PD (NS) | SCID-I | NS | NS |
| Taskapan et al. (2005) | Turkey | Cross-sectional | 40 | 48.3 | 62.5 | Stage 5D | HD (NS) | PRIME-MD | NS | GAD |
| Kokoszka et al. (2016) | Poland | Cross-sectional | 107 | 56.6 | 52.3 | Stage 5D | HD (NS) | MINI | NS | GAD, OCD, Panic disorder, PTSD, Social phobia |
| Tagay et al. (2007) | Germany | Cross-sectional | 144 | 63.1 | 50.7 | Stage 5D | NS | IES-R | Self-administered | PTSD |
| Preljevic et al. (2011) | Norway | Cross-sectional | 109 | 57.8 | 69.7 | Stage 5D | HD or PD (NS) | SCID-I | Psychiatrist | Panic disorder, Social phobia |
| Aghanwa and Morakinyo (1997) | Nigeria | Cross-sectional | 20 | 37.7 | 75.0 | Stage 5D | HD (NS) | PSE | Trained investigators | GAD |
| Filali et al. (2017) | Morocco | Cross-sectional | 103 | 49.7 | 54.4 | Stage 5D | HD ($\geq 1$ month) | MINI (Moroccan dialect) | Physician | Agoraphobia, GAD, Panic disorder, Social phobia |
| Cukor et al. (2007) | USA | Cross-sectional | 70 | 53.3 | 47.1 | Stage 5D | HD (NS) | SCID-I | NS | Agoraphobia, GAD, OCD, PTSD, Social phobia |
| Edmondson et al. (2013) | USA | Prospective, cohort | 388 | 55.1 | 51.3 | Stage 5D | HD (NS) | PCL-17 | Trained telephone interviewers | PTSD |
| **Anxiety symptoms** |  |  |  |  |  |  |  |  |  |  |
| **Asia** |  |  |  |  |  |  |  |  |  |  |
| Lai et al. (2005) | China | Cross-sectional | 167 | 60.7 | 47.9 | Stage 5D | PD ($\geq 2$ months) | HADS | Investigators | NA |
| Hou et al. (2014) | China | Cross-sectional | 81 | 51.1 | 64.2 | Stage 5D | HD ($\geq 1$ month) | SAS | Self-administered | NA |
| Wang et al. (2015) | China | Cross-sectional | 187 | 54.3 | 59.4 | Stage 5D | HD ($\geq 3$ months) | HADS | Self-administered | NA |
| Vasilopoulou et al. (2016) | Greece | Cross-sectional | 395 | NS | 56.2 | Stage 5D | HD (NS) | HADS | NS | NA |
| Gerogianni et al. (2019) | Greece | Cross-sectional | 414 | 63.5 | 63.3 | Stage 5D | HD (3 years) | HADS | NS | NA |
| Mok et al. (2019) | Hong Kong | Prospective, cohort | 182 | 57.8 | 57.7 | Stage 5D | PD (NS) | HADS | Self-administered | NA |
| Kohli et al. (2011) | India | Prospective, cohort | 30 | 42.4 | NS | Stage 5D | HD (NS) | STAI | Self-administered | NA |
| Aggarwal et al. (2017) | India | Cross-sectional | 200 | 50.1 | 64.0 | Stage 3-5D | NS | HADS | NS | NA |
| Marthoenis et al. (2020) | Indonesia | Cross-sectional | 213 | 47 | 65.3 | Stage 5D | HD (2 years) | HADS | NS | NA |
| Najafi et al. (2016) | Iran | Cross-sectional | 127 | 55.7 | 57.5 | Stage 5D | HD ($\geq 3$ months) | HADS | Self-administered | NA |
| Takaki et al. (2003) | Japan | Cross-sectional | 453 | 60.2 | 64.9 | Stage 5D | HD ($> 1$ year) | HADS | Self-administered | NA |
| Karasneh et al. (2020) | Jordan | Cross-sectional | 620 | 50.9 | 59.8 | Stage 5D | HD (NS) | CKD-SBI | NS | NA |
| Alshraifeen et al. (2020) | Jordan | Cross-sectional | 202 | 47.8 | 63.4 | Stage 5D | HD (NS) | GAD-7 | NS | NA |
| Macaron et al. (2014) | Lebanon | Cross-sectional | 51 | 64 | 60 | Stage 5D | NS | HADS | Investigators | NA |
| Semaan et al. (2018) | Lebanon | Cross-sectional | 83 | 67.9 | 60.2 | Stage 5D | HD ($\geq 1$ month) | HADS | Self-administered | NA |
| Bujang et al. (2015) | Malaysia | Cross-sectional | 1332 | 54.4 | 51 | Stage 5D | HD or PD ($\geq 3$ months) | DASS | NS | NA |
| Khan & Ahmad (2012) | Pakistan | Cross-sectional | 41 | 54.9 | 63.4 | Stage 5D | HD (NS) | HADS - Urdu version | Self-administered | NA |
| Shafi et al. (2017) | Pakistan | Cross-sectional | 156 | 47.3 | 61.5 | Stage 3 – 5D | HD ($\geq 9$ months) | HADS | Clinician | NA |
| Turkistani et al. (2014) | Saudi Arabia | Cross-sectional | 270 | NS | 58.2 | Stage 5D | HD ($\geq 6$ months) | HADS | Investigators | NA |
| Mosleh et al. (2020) | Saudi Arabia | Cross-sectional | 122 | 51.5 | 43.4 | Stage 5D | HD (3 years) | HADS | NS | NA |
| Lee et al. (2013) | South Korea | Cross-sectional | 208 | 58.9 | 61.1 | Stage 3-5 | NA | HADS | Self-administered | NA |
| Ng et al. (2015) | Singapore | Prospective, cohort | 159 | 53.6 | 60 | Stage 5D | HD ($\geq 1$ month) | HADS | Self-administered or Structured interview | NA |
| Griva et al. (2016) | Singapore | Prospective, cohort | 201 | 58.9 | 45 | Stage 5D | PD ($\geq 3$ months) | HADS | Self-administered | NA |
| Yoong et al. (2017) | Singapore | Cross-sectional | 526 | 56.1 | 58.7 | Stage 5D | HD ($\geq 3$ months) | HADS | Self-administered | NA |
| Rodrigo et al. (2013) | Sri Lanka | Cross-sectional | 100 | NS | 65 | Stage 1 - 5D | NS | Online 10-item validated anxiety screening tool (All Psych online) | Self-administered | NA |
| Ugurlu et al. (2012) | Turkey | Cross-sectional | 43 | 56 | 65.1 | Stage 5D | HD (≥ 5 months) | BAI | Self-administered | NA |
| Baykan and Yargic (2012) | Turkey | Cross-sectional | 83 | 49.1 (HD)<br>40.6 (PD) | 45.2 (HD)<br>41.5 (PD) | Stage 5D | HD or PD (≥1 year) | HADS | NS | NA |
| Cantekin et al. (2014) | Turkey | Cross-sectional | 120 | NS | 56.6 | Non-dialysis | NS | HADS | Investigator | NA |
| Dogu et al. (2018) | Turkey | Cross-sectional | 88 | 46.6 | 6.8 | Stage 5D | HD (≥ 6 months) | HADS | NS | NA |
| **Europe** |  |  |  |  |  |  |  |  |  |  |
| Bossola et al. (2010) | Italy | Cross-sectional | 80 | 64.2 | 63.7 | Stage 5D | HD (≥ 6 months) | HARS | Clinician | NA |
| Touil et al. (2019) | Morocco | Cross-sectional | 70 | 51.7 | 42.9 | Stage 5D | HD (5.2 years) | HADS | Interviewer | NA |
| Loosman et al. (2015) | Netherlands | Prospective, cohort | 100 | 67.9 | 57 | Stage 3-5 | NS | BAI | Self-administered | NA |
| Haverkamp et al. (2016) | Netherlands | Prospective, cohort | 249 | 58.9 | 61 | Stage 5D | HD or PD (≥ 6 months) | BAI | Self-administered | NA |
| Loosman et al. (2018) | Netherlands | Cross-sectional | 494 | 64.2 | 60 | Stage 5D | ≥ 3 months of dialysis | BAI | Self-administered | NA |
| Schouten et al. (2019) | Netherlands | Observational Prospective cohort | 508 | 65 | 62 | Stage 5D | HD or PD ($\geq 3$ months) | BAI | Self-administered | NA |
| Dumitrescu et al. (2009) | Norway | Cross-sectional | 161 | 53.9 | 54 | Stage 5D | HD (NS) | Purpose-designed self-report questionnaire | Self-administered | NA |
| Livesley et al. (1981) | Scotland | Cross-sectional | 85 | 43.3 | 65.9 | Stage 5D | HD ($\geq 2$ months) | GHQ & MHQ | NS | NA |
| Arenas et al. (2007) | Spain | Cross-sectional | 75 | 49.2 | 66.7 | Stage 5D | HD ($> 6$ months) | HARS | Clinician | NA |
| Pérez-Domínguez et al. (2012) | Spain | Cross-sectional | 59 | NS | NS | Stage 1-5D | HD (NS) | HADS | NS | NA |
| Rebollo Rubio et al. (2017) | Spain | Cross-sectional | 139 | 62.5 | 71.7 | Stage 5D | NS | HADS | Self-administered | NA |
| **North America** |  |  |  |  |  |  |  |  |  |  |
| Collister et al. (2019) | Canada | Cross-sectional | 50 | 64 | 52 | Stage 5D | HD (NS) | HADS | Self-administered | NA |
| Johnson and Dwyer (2008) | USA | Cross-sectional | 103 | NS | 62.1 | Stage 5D | HD (NS) | BAI | Self-administered | NA |
| Zhang et al. (2014) | USA | Cross-sectional | 72 | 52.3 | 68.1 | Stage 5D | HD ($\geq 6$ months) | BAI & HADS | NS | NA |
| Kutner et al. (1985) | USA | Cross-sectional | 127 | NS | 64.1 | Stage 5D | HD or PD (NS) | SAS | Self-administered | NA |
| Kopple et al. (2017) | USA | Cross-sectional | 246 | 56.8 | 58 | Stage 5D | HD ($\geq 6$ months) | BAI | Self-administered | NA |
| Feroze U et al. (2012) | USA | Cross-sectional | 170 | 56 | 54 | Stage 5D | HD or PD ($\geq 6$ months) | BAI | Self-administered | NA |
| **South America** |  |  |  |  |  |  |  |  |  |  |
| Stasiak et al. (2014) | Brazil | Cross-sectional | 155 | 55 (HD)<br>56.5 (PD) | 54.7 (HD)<br>44.4 (PD) | Stage 5D | NS | BAI & HADS | Investigators | NA |
| Palmieri et al. (2017) | Brazil | Cross-sectional | 170 | 56.2 | 59.4 | Stage 5D | HD (≥ 3 months) | HADS | Investigators | NA |
| Ottaviani et al. (2016) | Brazil | Cross-sectional | 100 | 52.3 | 66 | Stage 5D | HD (NS) | HADS | Self-administered | NA |
| Brito et al. (2019) | Brazil | Cross-sectional | 130 | 54.5 | 53.1 | Stage 5D | HD or PD (120 months) | BAI | Self-administered | NA |
| Galain et al. (2018) | Uruguay | Prospective, observational | 493 | 60.9 | 56.4 | Stage 5D | HD or PD (46-68 months) | HADS | Self-administered | NA |
| **Australia** |  |  |  |  |  |  |  |  |  |  |
| McKercher et al. (2013) | Australia | Prospective, cohort | 49 | 72.6 | 63 | Stage 4 | NS | BAI | Self-administered | NA |

**Abbreviations:** BAI - Beck's Anxiety Inventory; CKD-SBI – CKD Symptom Burden Index; DASS - Depression Anxiety and Stress Scale; GAD - Generalized Anxiety Disorder; GAD-7 – General Anxiety Disorder Scale GHQ - General Health Questionnaire; HADS - Hospital Anxiety Depression Scale; HARS - Hamilton Anxiety Rating Scale; HD - Hemodialysis; IES-R - Impact of Event Scale – Revised; OCD - Obsessive Compulsive Disorder; MHQ - Middlesex Hospital Questionnaire; MINI - Mini-international Neuropsychiatric Interview; NA – Not applicable; NS - Not specified; PCL-17, 17-item PTSD Checklist; PD - Peritoneal dialysis; PRIME-MD - Primary Care Evaluation of Mental Disorders; PSE - Present State Examination; PTSD - Post-traumatic Stress Disorder; SAS - Self-rating Anxiety Scale; SCID-I - Structured Clinical Interview for DSM-IV Axis I Disorders; SCL-90 - Symptom Checklist-90 Subscales of Depression and Anxiety; STAI, State-Trait Anxiety Inventory; Stage 5D – Stage 5 (Dialysis)

### Characteristics of patients in studies which examined anxiety disorders

A total of nine studies (14.8%) examined anxiety disorders among CKD patients (Table 1) (22–30). Five studies were conducted in Europe (n = 3, 33.3%) (25–27) and North America (n = 2, 22.2%) (22, 23), and the remainder in Asia (n = 2, 22.2%) (28, 29) and Africa (n = 2, 22.2%) (24, 30). The overall average patient age ranged from 37.6 to 63.1 years old (Table 1). All studies recruited ≤500 patients, of which 4 (44.4%) studies included ≤100 patients (22, 28–30). All studies included only dialysis patients while GAD was the most studied anxiety disorder (n = 5, 55.6%). The Structured Clinical Interview for DSM-IV Axis I Disorders (SCID-I) was the most commonly used instrument for diagnosing anxiety disorders (n = 3; 33.3%).

### Characteristics of patients in studies which examined anxiety symptoms

Fifty-two (85.2%) studies evaluated anxiety symptoms (Table 1) (8, 10, 13, 31–80). Most studies were conducted in Asia (n = 29, 55.8%) (8, 31, 33, 35–37, 40, 42, 44, 46, 49, 52, 54–56, 60–62, 64–66, 68, 69, 71–75), Europe (n = 11, 21.2%) (10, 13, 32, 34, 38, 41, 50, 51, 59, 67, 77) and North America (n = 5, 11.9%) (39, 43, 47, 48, 70). The overall average patient age ranged from 40.6 to 72.6 years old (Table 1). Nineteen studies (36.5%) included ≤100 patients (13, 32–34, 42, 44, 46, 50, 52, 53, 57, 59–61, 66, 70, 76, 78), while only 1 (2.0%) study recruited > 1000 patients (35). Dialysis patients (n = 44, 84.6%) was the most studied patient population while four studies included pre-dialysis patients (7.7%). The most frequently used instruments for assessing anxiety symptoms were the Hospital Anxiety and Depression Scale (HADS) (n = 31, 59.6%) and Beck’s Anxiety Inventory (BAI) (n = 12, 23.1%).

### Prevalence of anxiety disorders and symptoms among CKD patients

The pooled prevalence of anxiety disorders involving 1071 patients was 18.9% (95% CI: 10.6–27.2%) (Figure 2). Regarding anxiety symptoms, the pooled prevalence across studies involving 10,739 patients was 42.8% (36.3–49.3%) (Figure 3). Across continents, prevalence of anxiety symptoms was highest in Europe (49.1%, 95% CI: 31.7–66.4%), Asia (43.7%; 95% CI: 35.1–52.4%) and North America (42.8%, 95% CI: 29.1–56.5%). Between studies that included only either pre-dialysis or dialysis CKD patients, prevalence of anxiety symptoms was comparable, at 30.5% (95% CI: 14.5–46.5%) and 42.1% (95% CI: 35.0– 49.1%) respectively (Figure 4). Across instruments used for anxiety symptoms assessment, the prevalence of anxiety symptoms was similar between studies that utilized the BAI and HADS [39.2% (95% CI: 22.0–56.3%) vs 37.3% (95% CI: 30.7–44.0%)] (Figure 5).

**Figure 2:**
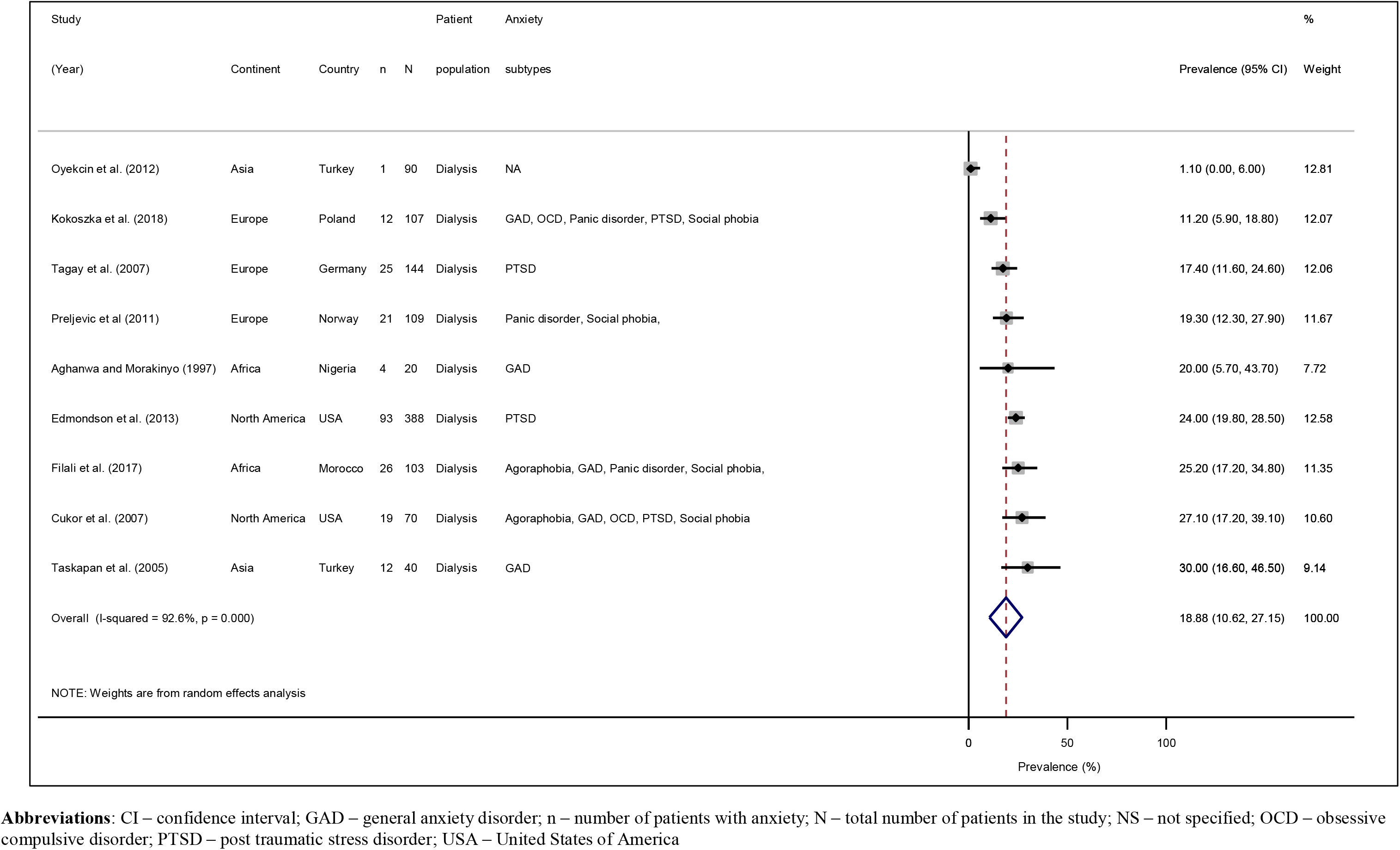
Prevalence of anxiety disorders among included studies (n = 9)

**Figure 3:**
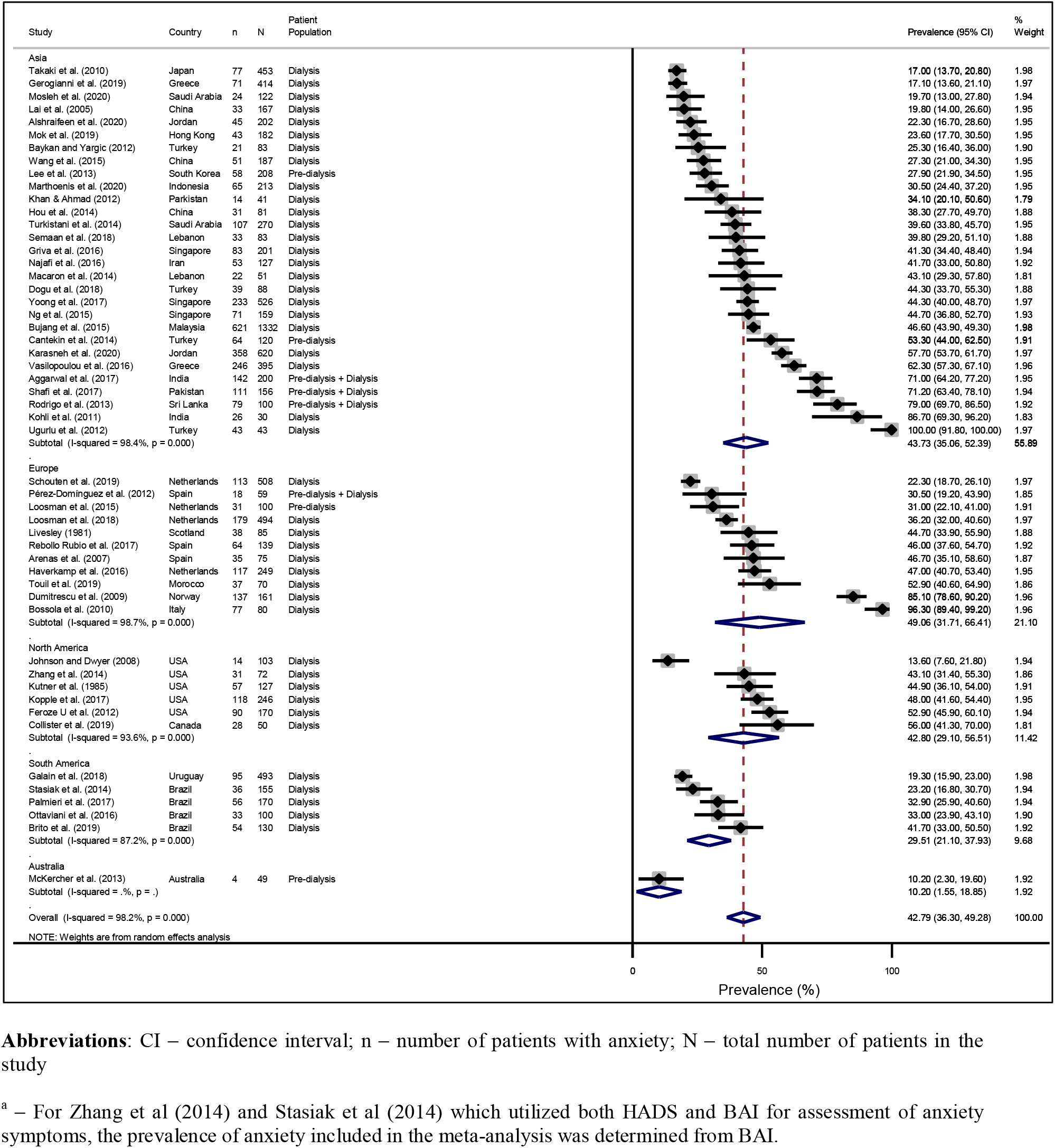
Prevalence of anxiety symptoms among included studies stratified by continent^a^

**Figure 4:**
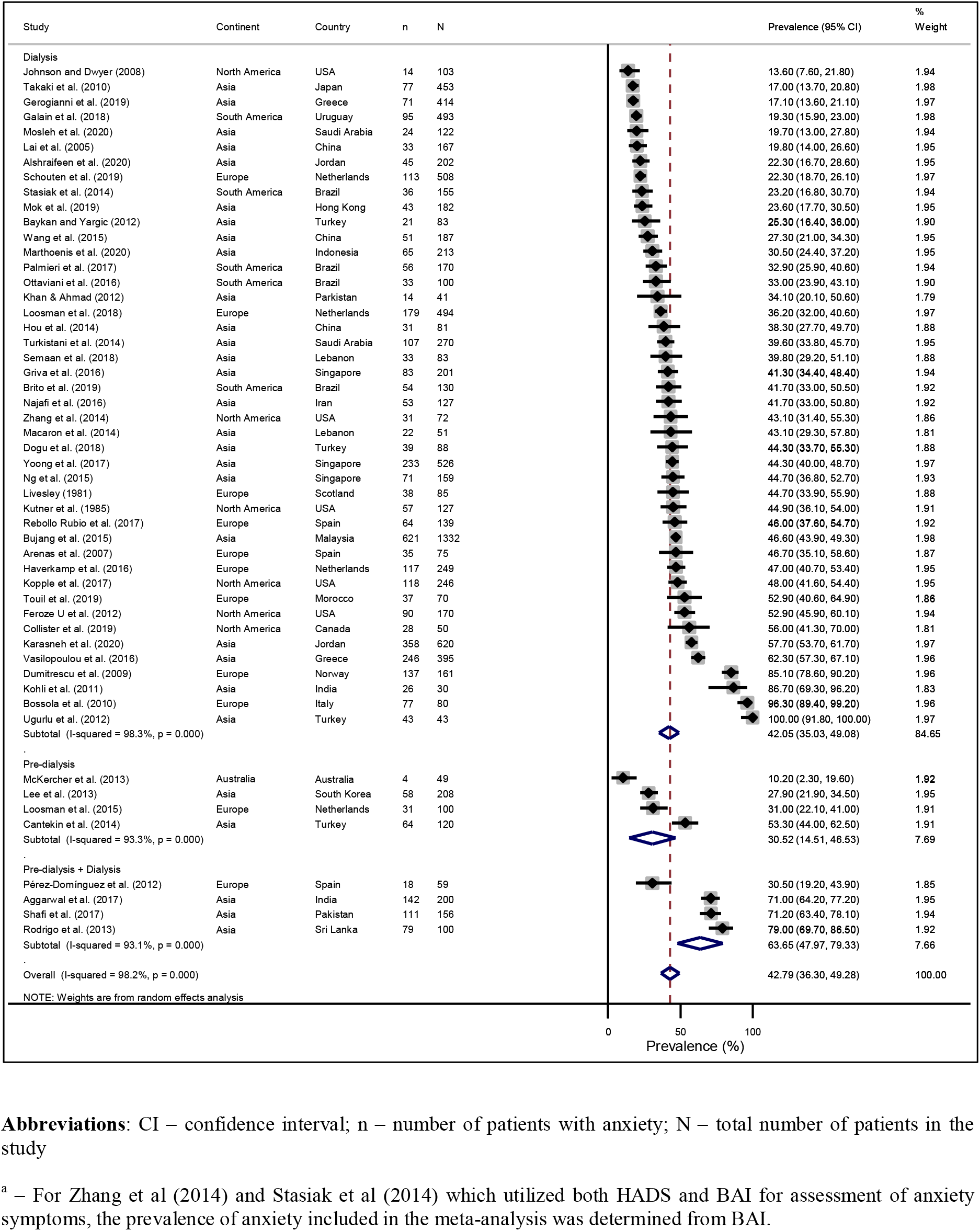
Prevalence of anxiety symptoms among included studies stratified by patient population^a^

**Figure 5:**
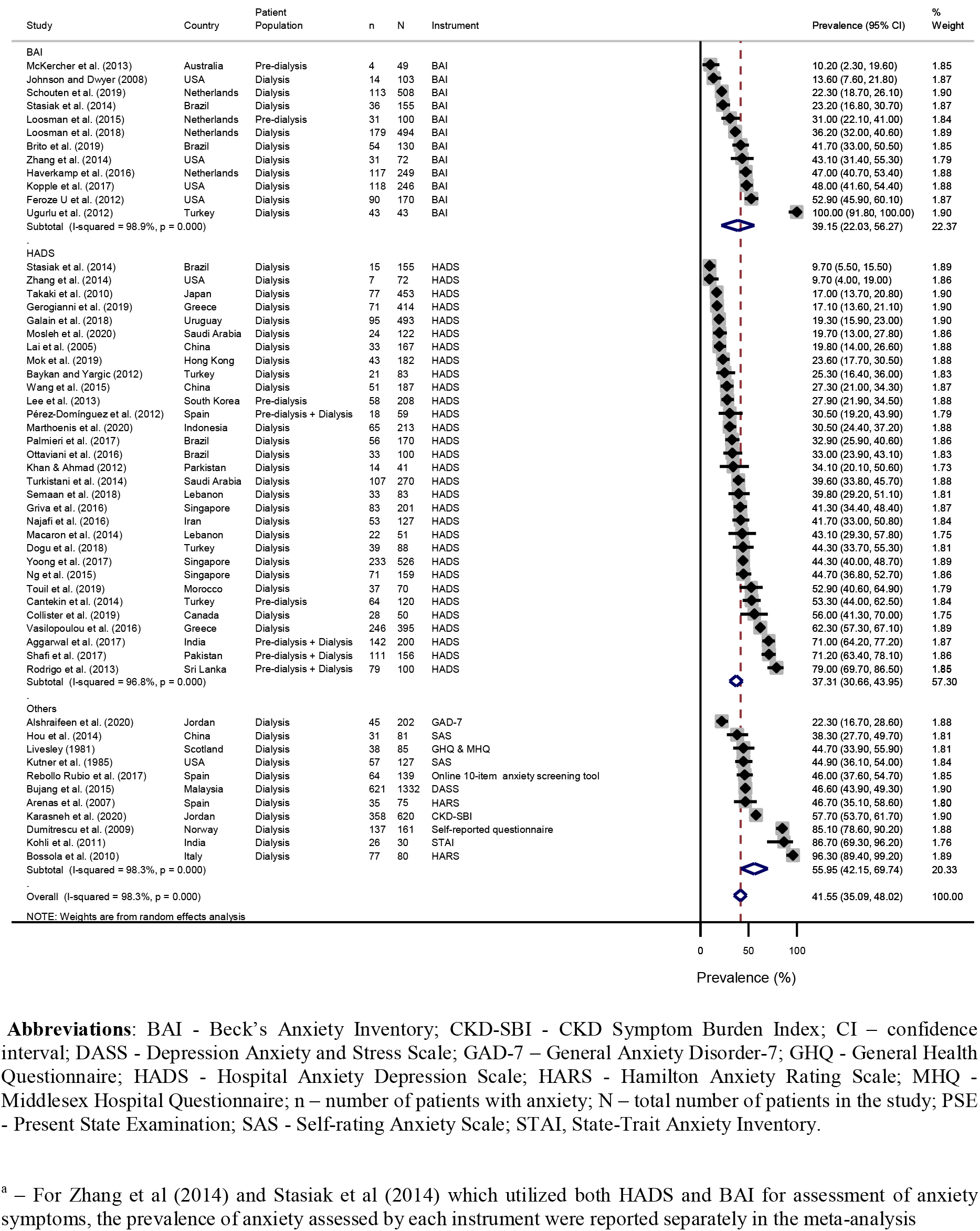
Prevalence of anxiety symptoms among included studies stratified by instrument used^a^

### Risk factors associated with anxiety symptoms and disorders among CKD patients

Thirty-six risk factors associated with anxiety were identified and categorized into four broad categories, namely patient-related, medical condition-related, therapy-related and psychosocial-related risk factors (Table 2).

**Table 2.** Risk factors associated with anxiety among chronic kidney disease patients

| **Risk Factors** | **Total number of supporting studies (Cited studies)** |
| --- | --- |
| **1) Patient-related factors** |  |
| *Patient demographics* |  |
| ↑ Age | 7 (24, 34, 36, 60, 65, 66, 76) |
| ↓ Age | 3 (54, 55, 63) |
| Male | 1 (66) |
| Female | 4 (27, 36, 42, 75) |
| Single marital status | 1 (66) |
| Urban residence | 1 (31) |
| Living with family members | 1 (61) |
| Ethnicity (Dark skin colour) | 1 (79) |
| *Socioeconomic status* |  |
| ↑ Unemployment | 1 (31) |
| ↓ Education level | 3 (31, 61, 74) |
| ↓ Income level | 1 (31) |
| *Dietary habits and level of physical activity* |  |
| ↑ Alcohol use | 1 (42) |
| ↓ Daily physical activity | 1 (70) |
| **2) Medical Condition-Related Factors** |  |
| *Psychiatric condition* |  |
| ↑ Depressive symptoms | 8 (29, 34, 42, 48, 56, 68, 74, 76) |
| ≥ 1 psychiatric disorder | 1 (26) |
| *CKD-related* |  |
| ↑ CKD severity | 1 (31) |
| ↑ CKD related symptoms (Cramps, weakness) | 1 (58) |
| ↑ Blood urea levels | 1 (31) |
| *CKD-related laboratory abnormalities* |  |
| ↓ Haemoglobin levels | 2 (10, 31) |
| ↓ Parathyroid levels | 2 (34, 55) |
| ↑ Serum phosphate levels | 1 (31) |
| ↓ Calcium levels | 1 (31) |
| *Dental-related conditions* |  |
| ↑ Dental decay | 1 (38) |
| ↑ Gingival overgrowth / redness / bleeding | 1 (38) |
| *Other factors* |  |
| ↑ Comorbidity index | 6 (8, 31, 34, 40, 74, 79) |
| Diabetes | 2 (54, 70) |
| ↑ Pain | 2 (24, 79) |
| ↑ Hospitalization admission/length | 2 (77, 79) |
| **3) Therapy-Related Factors** |  |
| *Medication-related factors* |  |
| Antidepressant use | 1 (63) |
| ↓ Medication compliance | 1 (49) |
| **4) Psychosocial Related Factors** |  |
| ↓ Perceived quality of life | 8 (8, 10, 32, 35, 53, 54, 57, 67) |
| ↑ Negative emotions | 1 (48) |
| ↑ Family problems | 1 (65) |
| ↓ Self-efficacy | 1 (64) |
| ↓ Social functioning | 1 (79) |
| ↓ Vitality levels | 2 (34, 79) |

**Abbreviations:** CKD – Chronic kidney disease; ↑ - increased; ↓ - decreased

Patient-related risk factors were divided into categories related to patient demographics, socioeconomic status, dietary habits, and level of physical activity. For patient demographic factors, increased age (24, 34, 36, 60, 65, 66, 76) and female gender (27, 36, 42, 75) were associated with anxiety while for socioeconomic factors, lower education (31, 61, 74) and income levels (31) were associated with increased risk for anxiety. Regarding dietary habits and physical activity, increased alcohol consumption (42) and reduced daily physical activity (70) were associated with a higher risk of anxiety.

Medical condition-related factors were divided into categories related to psychiatric disease-related factors, CKD condition-related, CKD-related laboratory abnormalities, dental related-conditions, and other factors. For psychiatric conditions-related factors, concomitant depressive symptoms (29, 34, 42, 48, 56, 68, 74, 76) and having more than one psychiatric condition (26) were associated with increased anxiety. Among CKD specific factors, greater severity of the disease (31) and presence of disease-related complications (58) were associated with increased anxiety. Regarding CKD-related laboratory abnormalities, lower hemoglobin (10, 31), hyperphosphatemia (31), hypocalcemia (31) and hypoparathyroidism (34, 55) were associated with anxiety. Additionally, dental disease (38) and a higher number of comorbidities (8, 31, 34, 40) were associated with anxiety.

For therapy-related factors, poorer medication compliance (49) and the use of antidepressants (63) were associated with increased anxiety.

Of the psychosocial-related risk factors in Table 2, poorer perceived quality of life (8, 10, 32, 35, 53, 54, 57, 67) was the most studied risk factors associated with anxiety. Increased negative emotions (48) and family problems (65) as well as reduced self-efficacy (64), vitality (34, 79) and social functioning (79) were associated with increased anxiety.

## Discussion

This review summarized the prevalence and risk factors associated with anxiety disorders and symptoms among CKD patients. To our knowledge, it is the first systematic review and meta-analysis examining the prevalence of anxiety disorders among CKD patients. Overall, the prevalence of anxiety disorders and symptoms among CKD patients were high, at 18.9% and 42.8% respectively.

For anxiety disorders, the prevalence among CKD patients was higher than that in the general population, which was estimated at 10.6% in a review by Somers et al. (81). In comparison to other patient populations with high disease burden, the prevalence of anxiety disorders was also higher than patients with malignancies (9.8%) (82), Type 2 diabetes mellitus (T2DM) (14%) (18) and, comparable with patients with stroke (3.8 – 25%) (83). Regarding anxiety symptoms, our results showed that the prevalence is higher than the findings from Murtagh et al. (2007) (15) which reported the prevalence of anxiety symptoms among CKD patients to be around 38%. Despite 12 years difference between the two studies and advancements made in the field of nephrology and the pathogenesis of renal diseases (84), anxiety remains a pervasive problem among CKD patients. When compared to other patient populations, prevalence of anxiety symptoms among CKD patients was higher than the general adult population (3.8–25%) (83) and long-term cancer survivors (21%) (85).

Compared to other neuropsychiatric disorders that CKD patients experience such as cognitive impairment and depression, the prevalence of anxiety disorders and symptoms were among the highest. A meta-analysis summarizing the prevalence of depression among CKD patients showed that 21.4 – 26.5 % of Stage 1–5 CKD patients had depressive symptoms (86), while the prevalence of CKD-related cognitive impairment among hemodialysis patients was between 30 to 60% (87).

In recent years, there is growing evidence suggesting the role of the “brain-renal axis”, which links the high prevalence of neuropsychiatric diseases such as anxiety among CKD patients, to the pathophysiology of CKD (88). Traditionally, anxiety is hypothesized to involve a complex interplay between psychological factors (e.g. excessive generalization of conditioned fear, social and environmental factors, genetic factors) and neurobiological factors like over-activity of the limbic areas of the brain (89). In CKD, postulated mechanisms for increased susceptibility to anxiety may involve inflammatory processes secondary to uremic toxins, oxidative stress from increased cytokine production, micro-vascular damage to the brain and involvement of the renin-angiotensin system (88). CKD patients also face challenges such as the need to cope with stressors ranging from adhering to complex medication regimens, dietary/fluid restrictions, managing CKD-related complications and adjusting to a lifelong dialysis regime. Poor management of these medical-related stressors compounded with psychosocial issues associated with chronic diseases such as financial insecurity and poor social support (90) may lead to anxiety.

In our study, the prevalence of anxiety symptoms among CKD patients was highest in Europe and Asia, as compared to studies performed in South America, Australia and Africa. Potential reasons for these differences could be due to impact of global economic crises and cross-cultural variations in disease perception. Healthcare financing across European countries operate via a mix of governmental funding, public and private healthcare insurances and copayments by patients (91). Following the 2018 global financial crisis, the World Health Organization reported substantial reduction in healthcare spending across 44 European countries, on a backdrop of increased unemployment (91). Importantly, Europe’s recovery from the crisis was slow compared to its peers due to the inadequate implementation of strict fiscal stimulus programs (91). For patients who are dependent on publicly funded health services, these added financial pressures may culminate in their development of anxiety symptoms.

Within Asia, multiple variants of anxiety-related distress syndromes which are closely linked to impaired cognitions about anxiety-related somatic and psychological symptoms exist. An example is the concept of “neurasthenia” (termed ‘shenjingshuairuo’) in China, which encompasses symptoms of excessive worry, headache and fatigue (92). Such cross-cultural variations in illness perceptions by patients are well-recognized and can influence the development of anxiety symptoms and disorders (93). Within the same country, racial, ethnic and cultural factors can lead to different prevalence of anxiety across patient groups. A study conducted in the United States by Asnaani et al. showed that Asian Americans had a lower likelihood of endorsing anxiety symptomology of all subtypes as compared to White Americans, which remained consistent after controlling for socio-demographic variables (94). Although the mechanisms for inter-ethnic differences in anxiety symptoms and disorders is unclear, ethnic identity could play a moderator role that mediates and maintains one’s psychological well-being during negative experiences (95). While cross-cultural variations in the study of anxiety symptoms and disorders are similarly expected to exist among CKD patients, few studies included in this review have examined these factors. There are also limited studies which have examined the prevalence of anxiety among patients in South America, Australia and Africa. Future studies involving larger patient populations are required to evaluate these differences and their role in anxiety disorders and symptoms among CKD patients.

Despite the high prevalence of anxiety and its implications on patient outcomes and quality of life, screening for anxiety symptoms and disorders among CKD patients has remained inadequately addressed in international nephrology guidelines (96). In the Kidney Disease: Improving Global Outcomes (KDIGO) 2012 guidelines, although a brief recommendation was made to address anxiety among CKD patients by involving a multidisciplinary team, no specific recommendations were made pertaining to the frequency of screening for anxiety symptoms, tools for assessing anxiety and the subgroups of patients to be screened. In our review, the prevalence of anxiety symptoms among dialysis patients was higher, albeit statistically non-significant when compared to pre-dialysis patients (44.5% vs 30.5%). Although this finding is limited by the small number of the studies examining pre-dialysis patients, the high prevalence of anxiety amongst dialysis patients highlights the need for screening of anxiety symptoms in this patient group.

Regarding anxiety screening tools, various instruments such as HADS, BAI and Generalized Anxiety Disorder 7 (GAD-7) have been utilized for evaluating anxiety symptoms among CKD patients. However, each instrument has inherent strengths and limitations. The main strength of HADS is its lower likelihood of being affected by confounding factors from physical symptoms due to the absence of somatic related questions (97). Furthermore, HADS has been validated for use among CKD patients (97). However, the usage of British colloquial expressions within the HADS limits the cross-cultural adaption of this tool (98). There is currently no consensus on the gold standard instrument for assessment of anxiety among CKD patients. Although the prevalence of anxiety assessed using the HADS and BAI were similar in our study, and they have comparable sensitivity and specificity for identifying anxiety (97, 99), the large number of questions within each questionnaire limits their applicability in daily clinical use. With limited healthcare resources, patient contact time and complexity of CKD related care in patient consultations, there has been growing interest in developing ultra-short screening questionnaires for anxiety symptoms in recent years. A 2019 study by Collister et al showed that the use of single question for anxiety screening in the Edmonton Symptom Assessment System had reasonable discrimination for anxiety among hemodialysis patients (78). Future studies are required for cross-cultural adaptation and validation of these screening instruments in larger studies and subgroups of CKD patients like pre-dialysis and peritoneal dialysis patients.

Regarding the frequency of anxiety screening, no defined optimal time-points for assessment exists. Hedayati et al. evaluated depression screening among CKD patients and suggestedscreening at interval events namely, the first consultation for CKD and when initiating dialysis for ESRD patients (100). For patients on dialysis, a recommendation was made for six-monthly interval screening during the first year of dialysis and yearly thereafter (100). Such a timeline may be adapted for screening of anxiety among CKD patients until an optimal screening frequency has been established.

Our study has also identified important risk factors associated with anxiety symptoms and disorders among CKD patients, which are categorised into patient-related, medical-condition related, therapy-related and psychosocial-related factors. Among patient-related factors, female gender, increasing age and lower educational levels were associated with increased risk of anxiety. The increased preponderance of anxiety among females concurs with findings in the general population (101), possibly due to an interplay of biological, genetic, cultural factors and clinical features such as increased rumination among women (102). For older patients, under-recognition of anxiety is important among CKD patients as anxiety symptoms share similarities with medical conditions such as hypothyroidism. Geriatric patients also have an increased tendency to somatise anxiety symptoms (103). Regarding education levels, individuals with higher educational levels may have higher resilience against stressors, thus having a protective effect against the development of anxiety (104). Among medical condition-related factors, concomitant depression was associated with increased risk for anxiety. A study by Tiller et al. showed that up to 90% of patients with anxiety have depression (105) and the two comorbidities were associated with increased risk of treatment failure and poor outcomes (106). In addition, anxiety and depression were recognized as bidirectional risk factors for predicting the occurrence of the other (107). Hence, physicians need to be cognizant of the risk factors associated with anxiety among CKD patients, permitting identification of early at-risk patients.

This review has identified important knowledge gaps in the literature regarding anxiety disorders and symptoms among CKD patients, which may guide and direct future research efforts. Future studies should consider (1) examining the prevalence of anxiety disorders and symptoms among pre-dialysis CKD and peritoneal dialysis patients; (2) assessing the prevalence of anxiety disorders and symptoms among CKD patients in South America, Australia and Africa; (3) developing time-efficient anxiety screening instruments for CKD patients; (4) determining optimal screening intervals for anxiety screening and (5) identifying subgroups of CKD patients who are at high risk for anxiety disorders or symptoms.

The findings from this review should be interpreted with several limitations. Firstly, the estimates for the prevalence of anxiety reported in the studies were unadjusted for age, and other patient demographics as the data required for the analyses could not be obtained. Nonetheless, with the increasing availability of electronic health records, multi-country collaborations and data-sharing, future meta-analyses may be performed to evaluate the adjusted prevalence rates of anxiety symptoms and disorders among CKD patients. Regarding the assessment of anxiety symptoms and disorders, significant heterogeneity exists due to the varying tools and criteria used for anxiety assessment and determination of diagnosis. While this is inevitable given the lack of recommendations from guidelines about the optimal tool for assessing anxiety among CKD patients, it is hoped that a standardized disease-specific anxiety instrument could be developed to allow more meaningful comparisons of the prevalence of anxiety across different patient populations. In this study, meta-analyses were not performed for the risk factors associated with anxiety symptoms or disorders due to relatively small number of studies available for each risk factor and significant heterogeneity in the assessment of these risk factors across studies. An evaluation of these risk factors should be considered in a future meta-analysis when more studies become available. In addition, while our search strategy encompasses key terms related to anxiety symptoms and disorders which have been used in other systematic reviews, the exclusion of articles that may be potentially relevant could not be ruled out. To minimize this, manual hand-searching of relevant articles was performed to enhance overall comprehensiveness.

## Conclusion

The overall prevalence of anxiety disorders and symptoms among pre-dialysis and dialysis CKD patients is high. Important risk factors associated with increased risk of anxiety symptoms and disorders include female gender, increased age, lower educational background, concomitant depression, increased comorbidities and a decreased perceived quality of life. Given the negative implications that anxiety symptoms and disorders have on health-related outcomes among CKD patients, there is a need to direct efforts into designing guidelines on screening anxiety symptoms and disorders among CKD patients. This will enable the early identification of at-risk patients for implementation of early intervention and treatment.

## Data Availability

All data is presented in the paper and the accompanying supplementary data files.

## Authors’ Contributions

JJB Seng was the study’s principal investigator and was responsible for the conception, literature review and design of the study. JJB Seng, CW Huang, PH Wee, LL Low, YLA Koong, H Htay, Q Fan and, WYM Foo the co-investigators. CW Huang, PH Wee and JJB Seng were responsible for the screening and inclusion of articles and data extraction. All authors contributed to the data analyses and interpretation of data. CW Huang, PH Wee and JJB prepared the initial draft of the manuscript. All authors revised the draft critically for important intellectual content and agreed to the final submission.

## Acknowledgements

Part of this work has been accepted as a poster abstract in the upcoming 57th ERA-EDTA Congress 2020.

## Guarantor’s name

JJB Seng is the guarantor of this work and, as such, had full access to all the data in the study and takes responsibility for the integrity of the data and the accuracy of the data analysis.

## Ethics approval

This study was exempted from institutional ethics board approval as it is a systematic review of existing literature.

## Availability of data and materials

The data and materials used in the study are available from the corresponding author on reasonable request.

